# Randomised trial of *Aureobasidium pullulans*-produced beta 1,3-1,6-glucans in patients with Duchenne muscular dystrophy: Favourable changes in gut microbiota and clinical outcomes indicating their potential in epigenetic manipulation

**DOI:** 10.1101/2022.12.09.22283273

**Authors:** Kadalraja Raghavan, Vidyasagar Devaprasad Dedeepiya, Naoki Yamamoto, Nobunao Ikewaki, Masaru Iwasaki, Ashwamed Dinassing, Rajappa Senthilkumar, Senthilkumar Preethy, Samuel JK Abraham

## Abstract

**Objective:** Duchenne muscular dystrophy (DMD) is an X-linked neuromuscular disorder that leads to increasing muscle weakening and early death. Steroids, the standard treatment of choice in slowing down disease progression, are plagued with adverse effects. Following anti-inflammatory and anti-fibrotic outcomes of an *Aureobasidium pullulans* strain N-163-produced beta 1,3-1,6-glucan food supplement in clinical and pre-clinical studies of DMD, herein we report their implications on the gut microbiome in patients with DMD.

**Design:** Twenty-seven patients with DMD were included in the pilot study (Control [n=9], N-163 [n=18]) which had previously reported the clinical decrease in inflammatory and fibrosis biomarkers. For the current study, whole genome metagenomic sequencing was performed in pre- and post N-163 intervention faecal samples of each of these participants.

**Results:** After N-163 beta-glucan administration, the constitution of the gut microbiome in all the participants was modified to one with positive outcomes on health. There was an increase in butyrate-producing species such as *Roseburia* and *Faecalibacterium prausnitzii*. There was a decrease in harmful bacteria associated with inflammation such as enterobacteria and *Alistipes*.

**Conclusion:** Beneficial reconstitution of the gut microbiome after N-163 beta-glucan administration, in addition to their implications in anti-inflammatory and anti-fibrotic outcomes, require further in-depth exploration on their roles in epigenetic manipulation.

## INTRODUCTION

Duchenne muscular dystrophy (DMD) is a fatal juvenile hereditary muscular disorder which affects 1 in 3500 to 5000 new born males globally, characterised by wasting and deteriorating muscle strength that progresses to death in the second or third decade of life. The dystrophin gene on the X chromosome is mutated leading to little or no production of the protein dystrophin [1,2]. The dystrophin protein, produced in differentiated myofibers, forms the dystrophin-associated glycoprotein complex, which links the cytoskeleton of the myofibers to the extracellular matrix. Myofibers are incredibly vulnerable to injury in the absence of dystrophin, which results in numerous cycles of degeneration and regeneration that eventually cause increased inflammation, fibrosis, and a progressive loss of muscle mass and function [1,2].

Despite the side effects connected with their prolonged usage, glucocorticoids continue to be the only medication clearly demonstrated to reduce disease development. Five drugs, in addition to the standard-of-care glucocorticoids, have acquired regulatory approval in some jurisdictions [3]. Exon-skipping or viral-mediated gene restoration are the recent therapeutic methods used to restore dystrophin currently, with the exon skipping approach being most advanced in the process of clinical application. Exon skipping converts a DMD mutation to a Becker muscular dystrophy-type mutation by inducing alternative splicing that avoids defective exons. This procedure involves anti-sense oligonucleotides. Becker muscular dystrophy is a milder variant of DMD, characterised by protein products that are expressed at lower levels than in healthy muscle but at higher levels than in DMD and nevertheless retain some functionality [1]. One of the antisense oligonucleotides-mediated exon skipping therapies that targets exon 51 became the first of its class to be approved by the United States regulators. However, the exon 51 skipping applies to only 13%–14% of patients with DMD. To allow for treatment of additional groups of patients, additional antisense oligonucleotides targeting other exons have to be developed and marketed [4]. In addition, there is a need for lifelong treatment. Gene therapy involves delivery of a mini dystrophin gene via viral vectors; however, the immunogenicity of the vector, off-target effects, and systemic body-wide delivery are major hurdles for both gene and exon-skipping therapies [5].

The regulation of host health is currently thought to be largely dependent on the gut microbiome. The complicated interactions between the host and many microbes are gradually being understood owing to the advent of molecular tools and methodologies (such as metagenomics, metabolomics, lipidomics, and metatranscriptomics). Currently, abnormalities in the gut microbiota (GM) are associated with a wide range of illnesses, including obesity, type 2 diabetes, hepatic steatosis, inflammatory bowel disorders, and various cancer types [6]. Weakness and muscle loss have been recently associated with GM dysbiosis and systemic inflammation. Notably, lower muscle mass and strength have been linked to greater circulating levels of tumour necrosis factor and interleukin (IL)-6 in older adults and people with inflammatory disorders. By regulating intestinal permeability, interorgan crosstalk, or directly targeting skeletal muscle, GM by-products such as short-chain fatty acids, phenolic products, bile acids produced by intestinal bacteria, and conjugated linoleic acid can enhance muscle glucose homeostasis, energy expenditure, protein synthesis, and physical performance [7]. Thus, the microbiome is likely linked as a potential major risk factor for neurological illnesses such as Alzheimer’s disease, autism spectrum disorder, multiple sclerosis, Parkinson’s disease, and stroke.

Nutritional supplements containing probiotics and prebiotics can aid in the recovery of the dsybiotic gut. With demonstrated effectiveness against metabolic illnesses, diabetes, cancer, cardiovascular diseases, and neurological diseases, beta-glucans are among the most promising dietary supplements. Beta-glucans produced by two strains of the black yeast *Aureobasidium pullulans*, AFO-202 and N-163, have been demonstrated to have promising results in the treatment of several illnesses and ailments [8-11]. The AFO-202 strain beta-glucan has been demonstrated to improve behaviour and sleep patterns in addition to raising levels of synuclein and melatonin in children with autism spectrum disorder [11]. Evaluation of the GM of individuals with autism spectrum disorder following consumption of AFO-202 beta 1,3-1,6-glucan in a randomised pilot clinical study demonstrated its role in the effective control of curli-producing Enterobacteriaceae that results in synuclein misfolding and accumulation, in addition to an increase in beneficial bacteria [12]. In another study, the beta-glucans produced by the AFO-202 and N-163 strains, increased gut microbial diversity, controlled pathogenic bacteria, promoted healthy bacteria, and induced beneficial changes in faecal metabolites individually and in combination in an animal model of NASH [13]. The findings of this study supported the use of AFO-202 beta-glucan as a metabolic regulator and N-163 beta-glucan as an immune modulator. Taken together, these substances can be regarded as potentially useful and safe adjuncts in the treatment of NASH as well as a preventative measure in other chronic inflammatory and immune-dysregulated conditions.

The current study aimed to evaluate the gut microbiome of participants included in a pilot study that was undertaken to evaluate the immunomodulatory efficacy of the *A. pullulans* strain N-163-produced beta 1-3,1,6-glucan in comparison with that of a conventional therapeutic regimen in patients with DMD [14]. The clinical results of the study showed that supplementation with the N-163 beta glucan food supplement produced disease-modifying beneficial effects: a significant decrease in inflammation and fibrosis markers such as IL-6, IL-13, and transforming growth factor-beta; increase in dystrophin; and improvement in muscle strength in DMD subjects over a period of 45 days.

## METHODS

The study was registered with the clinical trial registry of India, CTRI, ref no. CTRI/2021/05/033346. The study was approved by the Institutional Ethics Committee (IEC) of Saravana Multispeciality Hospital, India on 12 April 2021. The caregivers of each participant provided their informed consent for inclusion before participation in the study. The study was conducted in accordance with the Declaration of Helsinki.

This study involved individuals with DMD and was an investigator-initiated, single-centre, randomised, open-label, prospective, comparative, and two-arm study. The study was conducted over a period of 45 days. The two therapy modalities comprised the following:

Treatment arm I (control): Conventional treatment plan including 6–24 mg of T. deflocort (steroid), T. calcium, and vitamin D 1000 in addition to typical routine physiotherapy for joint mobility.

Treatment arm II: N-163 beta-glucan (16 mg gel) in one sachet once daily in addition to standard care.

The inclusion criteria were male participants aged 6 to 18 years who had a molecular diagnosis of DMD and provided written informed consent to participate in the trial. The exclusion criteria included patients who have recently (within the last month) or concurrently participated in any other therapeutic trial, have a known or suspected malignancy or any other chronic disease, or at the investigator’s discretion, have a clinically relevant limitation of renal, liver, or heart function.

Before the patients were enrolled in the trial, background data on gender, date of birth, age, eating habits, current medical history, medication, treatment, prior history, and allergies (to drugs and food) were gathered. In addition, functional foods, health foods, foods high in - glucans, and foods containing beta-glucans were also considered. Consumption of test foods, body temperature, intake of food for specific health purposes, functional and health foods, intake of restricted foods, subjective symptoms, visits to medical facilities, therapy, and usage of medications were all noted during the study.

Foods that were prohibited included beta-glucan-rich supplements including those formulated with concentrated beta-glucan that has been extracted from yeast, barley, mushrooms, and seaweed. Foods that claim to improve gut dysbiosis such as yogurt, lactobacillus beverages, bifidobacteria powder, propolis, and lactoferrin, were also excluded.

### Collection and preparation of faeces samples

Faeces samples were collected at baseline and 90 days following the intervention using a sterile faecal collection kit. The samples were stored at 20 °C until they were transported to the lab and processed. Samples for DNA extraction were kept at 80 °C until they were required for analysis.

### DNA extraction

Total microbial DNA was extracted from each sample faeces using the QIAamp DNA Mini Kit (Qiagen) according to the manufacturer’s instructions. Every group of samples was extracted using a negative buffer control (extraction control).

### Metagenome sequencing

A total of 27 faecal samples were sequenced with a HiSeqX system utilising 151 bp reads. The samples were collected for a metagenome study of the entire genomic. The readings were initially screened for contamination with human DNA. Around 20.51% of the human genome was aligned. Following filtering, the reads were aligned to the genomes of bacteria, fungi, viruses, and archaea. Around 51% of the bacterial genome was aligned overall. However, only 0.04% to 0.08% of the viral, fungal, and archaeal genomes were aligned.

The scaffolds were produced using a de novo assembly utilising the pre-processed reads. The prediction of genes was then performed using these scaffolds.

The following bioinformatics workflow was used to conduct a metagenomic study of whole genome sequencing. The adapters were removed once the raw data quality was examined. To exclude reads that were not aligned to the human genome, the low-quality reads were first assembled using the metaSPAdes de novo assembler for metagenomics. After assembly, Prodigal was used to carry out the gene prediction. Using the DIAMOND-MEGAN5 tool, the predicted genes were then compared to genes already present in the NCBI database. Shannon plots for microbial alpha diversity were produced. SEED-based functional assignment was performed to identify the functional differences between the control and treatment groups, followed by subsystem analysis using the linear discriminant analysis (LDA) effect size (LEfSe) method. Based on the taxonomic abundance in the provided samples, the occurrence of the dominant microbial community was investigated at multiple levels (phylum, class, order, family, and genus). Dominance was determined based on the quantity of sequences collected from samples, make-up of the community, and distribution of contig sizes. The analysis was screened to remove chimeric sequences.

### Statistical analysis

Origin Lab Origin 2021b software and the Microsoft Excel statistics package were used to analyse the statistical data. Statistical significance was set at *p* < 0.05. For metagenomic sequencing data, including taxonomy, non-parametric tests such the Friedman test for repeated-measures and Kruskal-Wallis test for independent measures were applied.

## RESULTS

Twenty-eight patients were screened and 27 were randomised to the control (n = 9) and treatment (n = 18) groups. One patient was disqualified owing to misrepresentation of diagnosis [14].

Regarding taxonomic profiling, in both the pre- and post-intervention, the bacterial kingdom was the most abundant. The phylum Firmicutes was the most abundant followed by Bacteroidetes. Most of the read genera belonged to *Bacteroides* followed by *Prevotella* (Figure 1). *Faecalibacterium prausnitzii* was the most abundant species (Figure 2).

**Figure 1:**
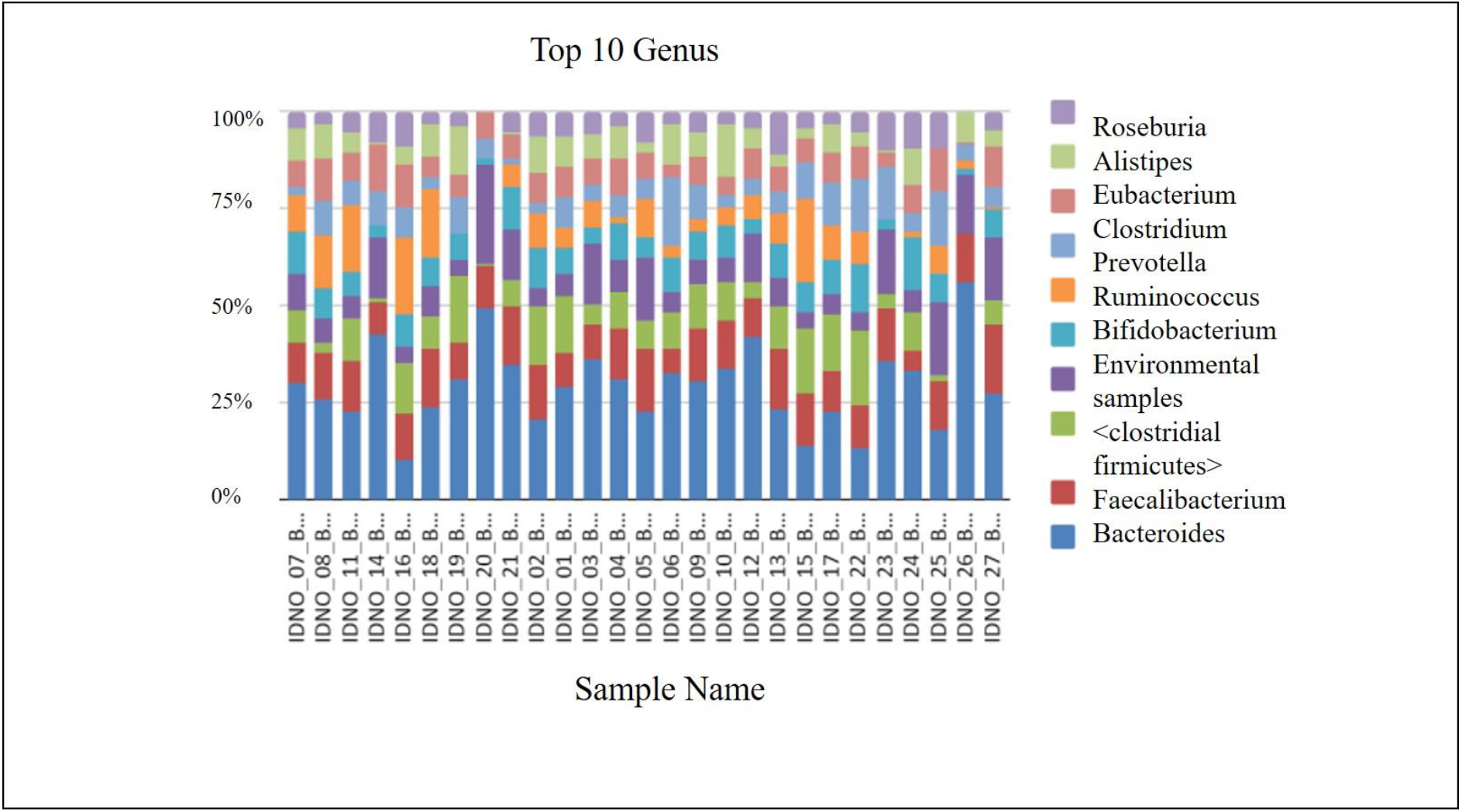
Distribution of genera post-N-163 beta-glucan intervention showing *Bacteroides* as the most abundant genus

**Figure 2:**
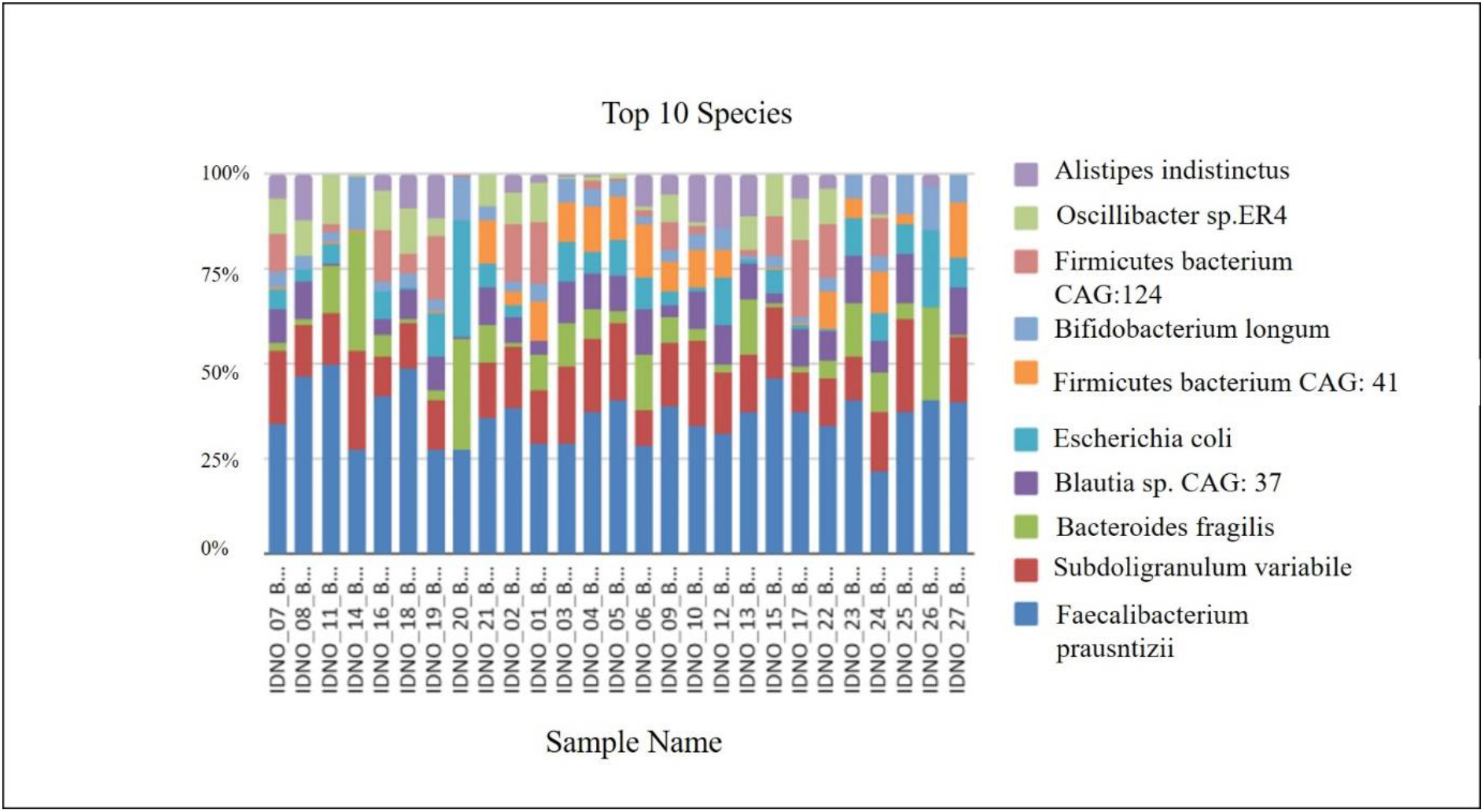
Distribution of species post-N-163 beta-glucan intervention showing *Faecalibacterium prausnitzii* as the most abundant species.

Analysis of individual taxa indicated a decrease in *Enterobacteriaceae* (*p* = 0.81), *Alistipes* (*p*= 0.81), *Akkermansia muciniphila* (*p*=0.97) and *firmicutes* (*p* =0.7) (Figure 3).

**Figure 3:**
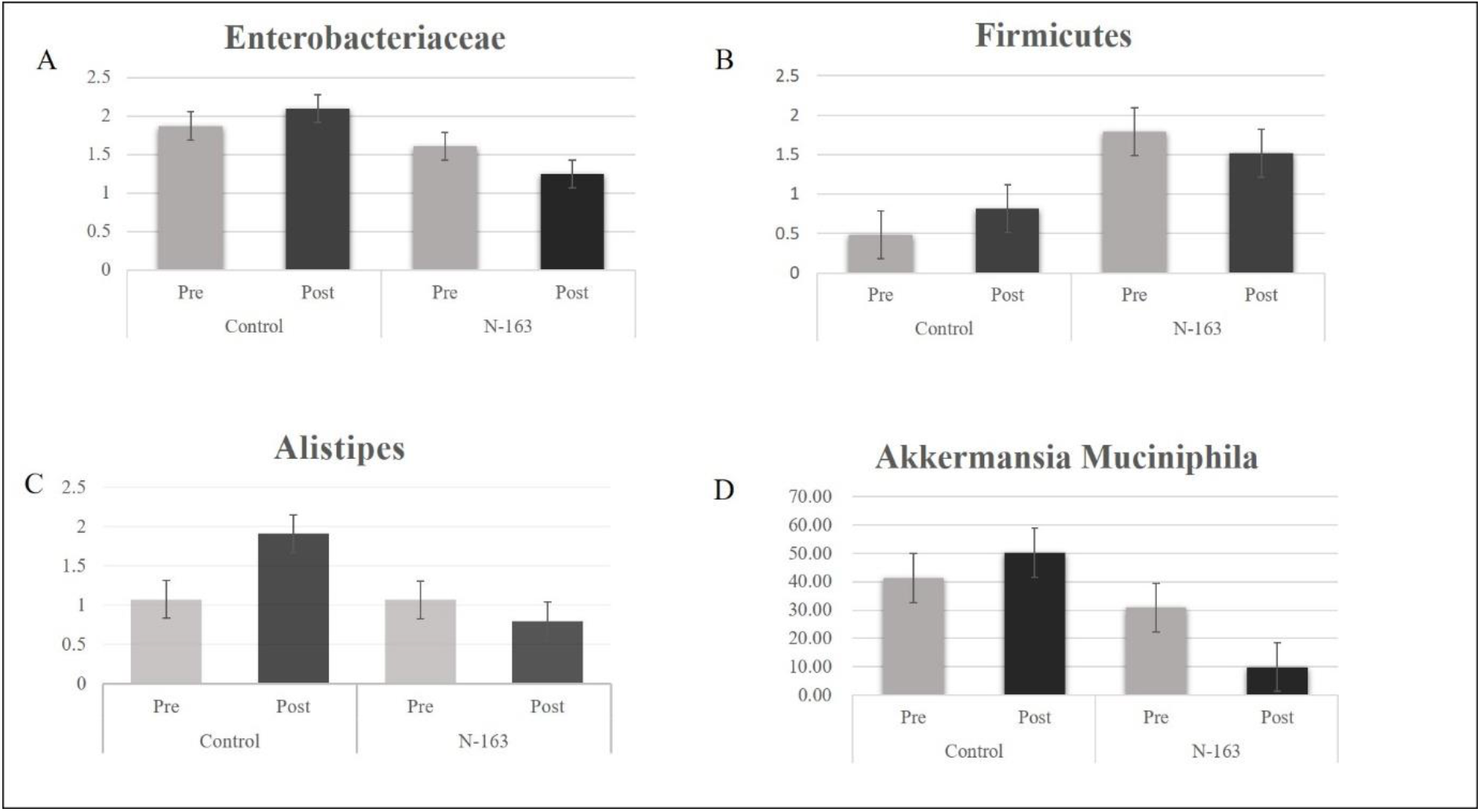
Decrease in abundance of *Enterobacteria, Firmicutes, Alistipes*, and *Akkermansia* post N-163 beta-glucan intervention. Error bars indicate standard error.

An increase in *Lactobacillus* (*p* = 0.14), *Roseburia* (*p* = 0.45), *Bifidobacterium* (*p* = 0.74), and *Prevotella* (*p* = 0.69) was observed (Figure 4).

**Figure 4:**
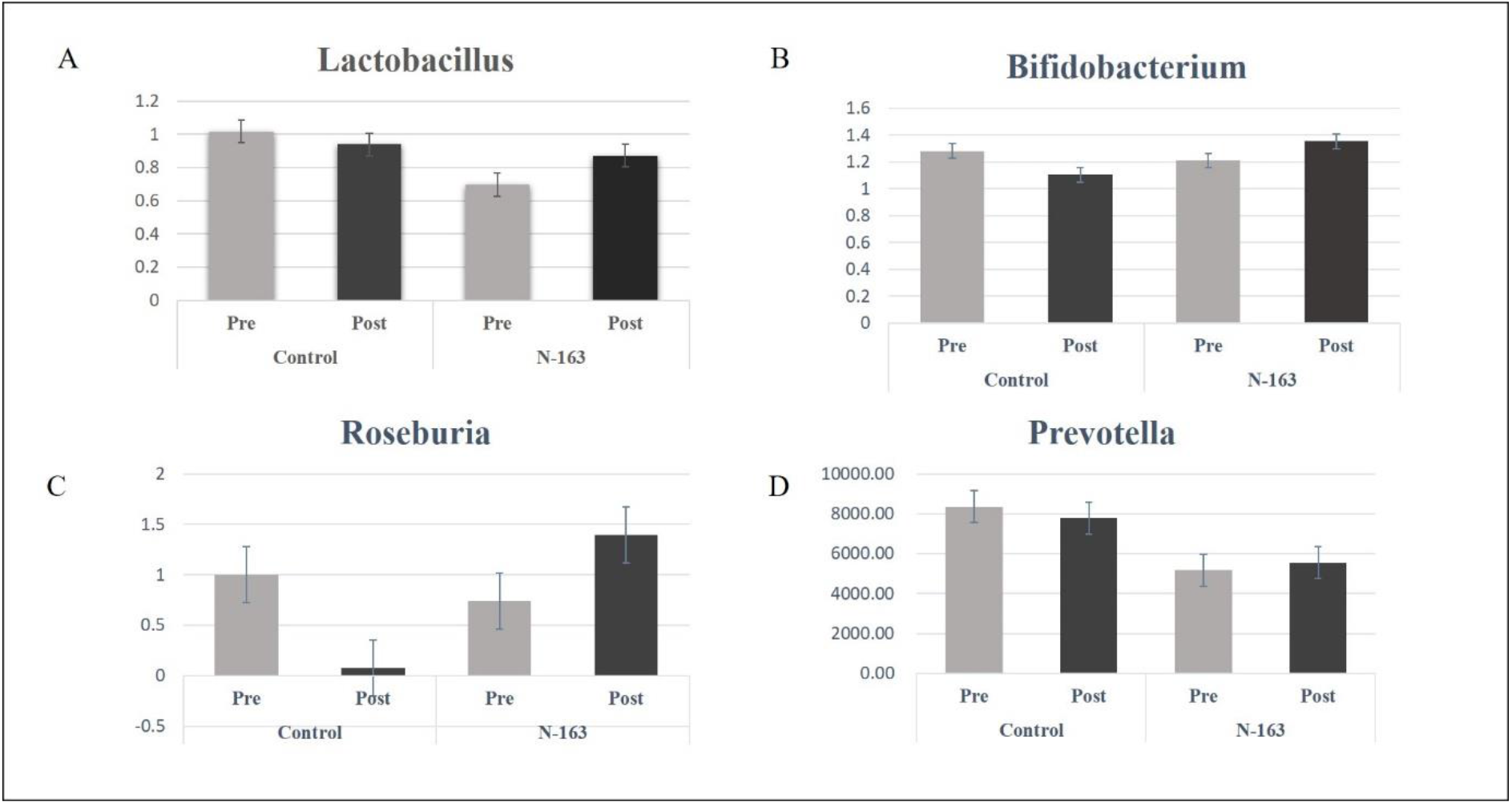
Increase in abundance of *Lactobacillus, Bifidobacterium, Roseburia*, and *Prevotella* post N-163 beta-glucan intervention. Error bars indicate standard error.

Results of the SEED-based functional assignment analysis showed an increase in carbohydrate fermentation, DNA repair, fatty acid metabolism, fructose utilisation, and protein biosynthesis by bacteria (Figure 5). The detailed results of the taxonomic and SEED analysis are presented in Supplementary File 1.

**Figure 5:**
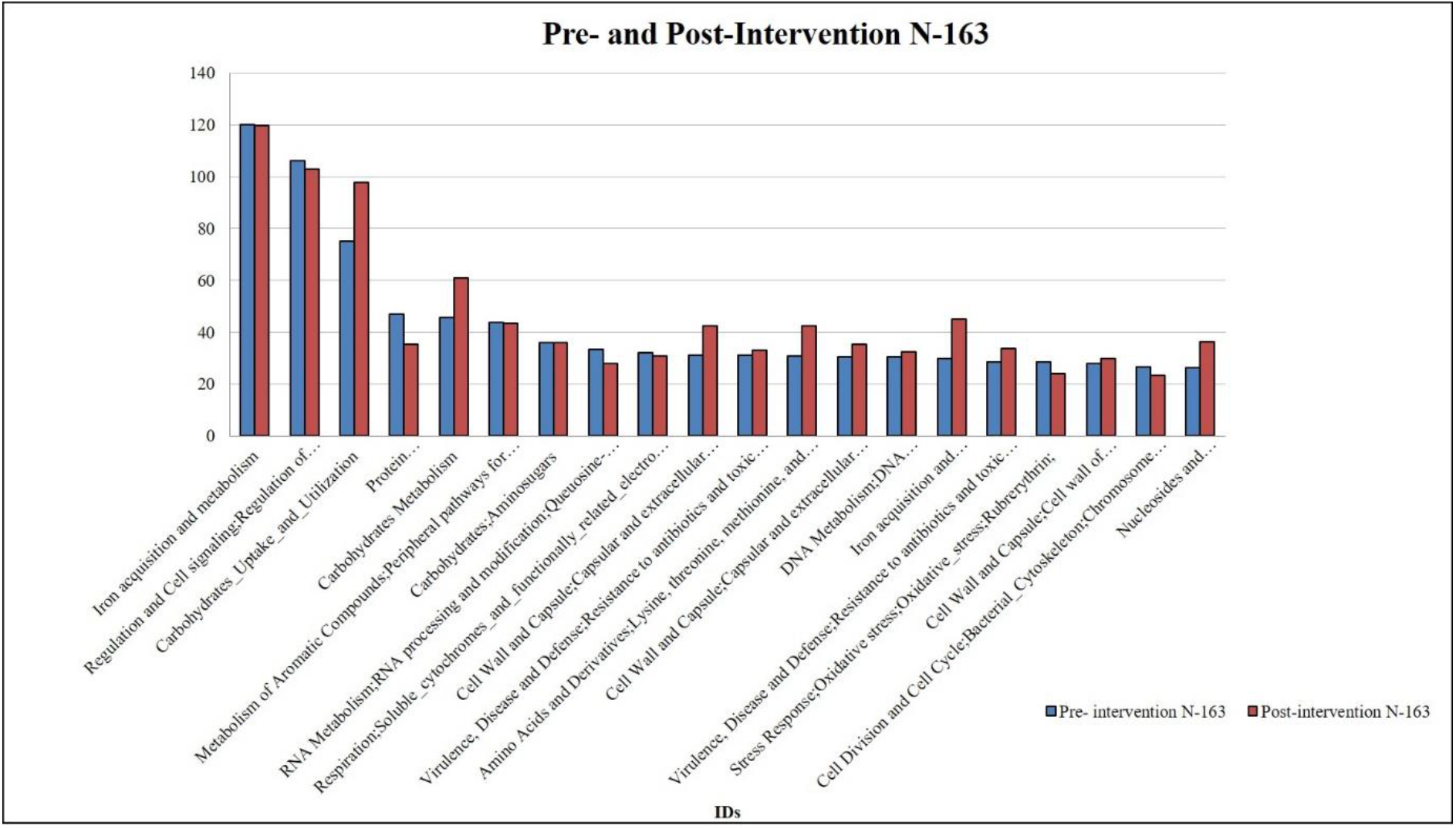
SEED-based functional analysis of metabolic and functional pathways

## DISCUSSION

The current disease modifying approaches [3,15] for DMD fall into two categories, viz., (i) Standard treatment such as corticosteroids and (ii) Supportive approaches including anti-inflammatory, anti-fibrotic agents and experimental approaches of muscle regeneration such as exon skipping therapies The standard treatment of corticosteroids enhances muscle strength and function in boys with DMD, according to evidence from randomised controlled trials. Although concerns persist about their capacity to prolong walking, when to begin treatment, the long-term balance of benefits versus risks, and the choice of corticosteroid or other regime remain a question[15]. Furthermore, whether some of the more severe adverse effects (such as excessive weight gain, cataracts, behavioural problems, delayed growth, and osteoporosis) outweigh the intended advantages remains unclear [16].

In the present study, the objective of using beta glucans is to serve as an adjunct support exerting anti-inflammatory and anti-fibrotic actions based on its earlier reports of decreasing anti-inflammatory markers such as C-reactive protein (CRP), Tumour necrosis factor-Alpha (TNF-α), IL-6 along with exerting anti-fibrotic effects in clinical and pre-clinical studies of COVID-19 [8] and Non-alcoholic steatohepatitis (NASH) [10]. Although the beneficial immune-regulatory and systemic effects of the A.pullulans produced beta-glucans by regulating the gut dysbiosis has been reported pre-clinically as well as in clinical studies of multiple sclerosis, autism spectrum disorder, and metabolic syndrome [6,12,13], the beneficial effects in a rare genetic disease such as DMD have not yet been reported. To our knowledge, this is the first clinical study to demonstrate the beneficial effects of the gut microbiome in DMD in addition to the previously reported correlation of anti-inflammatory and anti-fibrotic clinical outcomes [8-14].

### Anti-inflammatory and anti-fibrotic effects through the gut microbiome

*Faecalibacterium prausnitzii* is an important member of the phylum Firmicutes and one of the most prevalent bacteria in the normal human microbiota. Depletion of *F. prausnitzii* has been observed in patients with Crohn’s disease and other gastrointestinal illnesses. Strains of *F. prausnitzii* might make excellent candidates for next-generation probiotics [17].

High levels of butyrate or butyrate-producing taxa were noted in voluntary free wheel running mice and in voluntary running rats fed a 25% casein-sucrose diet. According to a study on humans, *F. prausnitzii* (order Clostridiales), which produces butyrate, benefits from exercise, and a decline in its population may be linked to high frailty. Intestinal abundance of *Bacteroides* spp. was similarly correlated with wheel running distance in mice and brisk walking in elderly women. *Bacteroides* spp. have been demonstrated to be inversely correlated with obesity associated with high-fat and high-carbohydrate diets and to a decline with age [17]. In the current study, post N-163 beta-glucan consumption, *F. prausnitzii* was the most prominent species and *Bacteroides* the most prominent genus. Lactic acid, a key GM metabolite precursor of short-chain fatty acids, is produced by the bacteria *Lactobacillus* (phylum Firmicutes, order Lactobacillales), *Streptococcus* (phylum Firmicutes, order Lactobacillales), and *Bifidobacterium* (phylum Actinobacteria, order Bifidobacteriales), which are present in probiotic yoghurts and commercial dietary supplements. By increasing the expression of tight junction proteins, certain species or strains of *Lactobacillus* or *Bifidobacterium* have been shown to favour the integrity of the intestinal barrier, as well as an increase in muscle weight and muscle fibre size [18]. In the present study, an increase in *Lactobacillus* and *Bifidobacterium* was observed in patients with DMD after N-163 supplementation which would be helpful to build muscle. The symbiotic microbiota of a host serves critical roles, and the loss of helpful microorganisms may encourage the growth of microbial pathobionts. In particular, the inflammatory response has been noted to be enhanced by the blooming of potentially dangerous Proteobacteria, particularly Enterobacteriaceae [19]. The current study observed a decrease in Enterobacteriaceae, which also correlates with the decrease in inflammatory markers such as IL-6 and tumour necrosis factor-α. In a previous study, the expression of atrophy markers was reduced when *Lactobacillus* species were restored through oral supplementation with *L. reuteri* 100-23 and *L. gasseri* 311476. This phenomenon was correlated with a decrease in inflammatory cytokines which has also been observed by supplementation with N-163 beta-glucan. *Roseburia* spp. break down food elements to promote their growth and metabolic processes. They are a type of commensal bacteria that produce butyrate and other short-chain fatty acids that have an effect on colonic motility and immunity and anti-inflammatory activity. *Roseburia* spp. may also act as probiotics to restore healthy flora or as biomarkers for pathologies with symptoms such as the formation of gallstones [20]. An increase in the abundance of *Roseburia* spp. was observed in the present study. A diet rich in vegetable fibres, prebiotics that support the de facto microbial diversity and a lower inflammatory profile, have been reported to favour the growth of *Prevotella* (phylum Bacteroidetes), a mucin degrader enriched in human enterotypes [20]. An increase in *Prevotella* was observed in the current study. *Alistipes* affects the epithelium and causes inflammation. Firmicutes, particularly those from the family Lachnospiraceae, may contribute to the development of depression by influencing the inflammatory response of the host. According to recent data, patients with PD and MS have been reported to exhibit an increase in the abundance of *Akkermansia* [21]. Therefore, a decrease in *Alistipes, Akkermansia*, and *Firmicutes* after N-163 beta glucan supplementation is beneficial.

### Potential of epigenetic regulation through the gut-microbiome

Among the supportive approaches to DMD, regeneration of lost muscle cells has been attempted. One such approach is to regenerate satellite stem cells and these satellite cells, especially their fate is modulated by epigenetic regulatory pathway involving mitogen-activated protein kinase (MAPK) p38γ/MAPK12 which mediates commitment during asymmetric division acting downstream of the dystrophin-associated glycoprotein complex and polarity establishment. These satellite cells, are essential for muscle regeneration after damage and are responsible for postnatal muscle growth. Dystrophin is expressed in satellite cells where it plays a crucial role in controlling the formation of satellite cell polarity and subsequently, effective asymmetric division in addition to its structural involvement in myofiber stability. Beta-glucans have been shown to increase plasma dystrophin in a pilot clinical study [14]. Therefore, the effects on dystrophin enhancement may also involve epigenetic pathways. For instance, a maternally inherited trans-generational epigenetic silencing has been shown to result in exon skipping and dystrophin rescue [22]. Gut microbiota and their metabolites are considered to be significant epigenetic regulators. Although the molecular processes by which the GM interacts with the host cells and controls gene expression are not fully understood, the presence of the interaction of a number of metabolites derived from microorganisms at the DNA, RNA, and histone levels has been demonstrated. By inducing epigenetic changes, including DNA methylation, histone modifications, and regulation by noncoding RNAs, the altered function and composition of the GM have been linked to influence the gut-brain axis [23,24].

Thus, the beta-glucans capable of influencing dystrophin expression and modulating gut-dysbiosis may have exerted the positive clinical anti-inflammatory and anti-fibrotic effects through epigenetic control via the gut microbiome which needs further evaluation.

Another major factor to be considered are the adverse effect associated with existing therapies for DMD. Immune-suppression related side-effects of standard therapies such as corticosteroids and off-target effects of gene-based therapies can be addressed by the use of adjuncts such as the beta-glucans by their capabilities of immune-regulation, as they have been shown to be potent inducers of epigenetic and functional reprogramming of innate immune cells, by a process called “trained immunity” [25]. The timing of administration of the different treatments for DMD while needs to be researched, the epigenetic effects being present from pre-natal state and the beta glucans being a safe food supplement even during pregnancy makes it be suggested for administration right from the age of diagnosis of DMD and even during maternal nutrition [26] for better treatment outcome.

The current study has some limitations. The extent of changes in inflammation and fibrosis may vary from patient to patient and it is imperative to investigate whether the more favourable changes in the GM was observed in patients with more pronounced reductions in inflammation and fibrosis. In addition, the amount of beta-glucan that reaches the intestinal tract after oral consumption should also be studied as this may vary from individual to individual and the effects exerted may also vary.

## CONCLUSION

Our study yielded a beneficial reconstitution of the gut microbiome after oral consumption of the biological response-modifier glucans which is correlated with biomarkers levels amounting to slowing down the progress of the pathology in DMD clinically [14], with a similar correlation in pre-clinical study [27] as well. These gut microbiome modulations, in addition to their implications in anti-inflammatory and anti-fibrotic outcomes, require further in-depth exploration of their roles in epigenetic manipulation. Extensive research on this safe approach may unravel further insights into their potential in other rare genetic diseases which don’t have a definitive intervention at this point of time.

## Data Availability

All data produced in the present work are contained in the manuscript

## Acknowledgements

The authors thank

1. The Government of Japan and the Prefectural Government of Yamanashi for a special loan and M/s Yamanashi Chuo Bank for processing the transactions.
2. Ms. Sunitha, Mr. Vincent, Mr. Shivakumar (Physiotherapist), Dr. Madhankumar and the staff of Kenmax & Sarvee Integra, for their assistance during the clinical study and data collection of the manuscript.
3. Fr. Francis Xavier, Fr. Vargheesh Antony and Fr. Marianathan of JAICARE for their support during the clinical study.
4. Ms. Eiko Amemiya of II Dept. of Surgery, University of Yamanashi for her secretarial assistance.
5. Mr. Yoshio Morozumi and Ms. Yoshiko Amikura of GN Corporation, Japan for their liaison assistance with the conduct of the study.
6. Loyola-ICAM College of Engineering and Technology (LICET) for their support to our research work.

